# Consistency of the continuous flow pressure gradient despite aortic arch anomalies co-existing with coarctation

**DOI:** 10.1101/2023.10.30.23297763

**Authors:** Arash Ghorbannia, Andrew D. Spearman, Shahd Sawalhi, Ronald K. Woods, Mehdi Maadooliat, John F. LaDisa

## Abstract

**Aims:** Severity assessment for coarctation of the aorta (CoA) is challenging due to concomitant morphological anomalies (complex CoA) and inaccurate Doppler-based indices. Promising diagnostic performance has been reported for the continuous flow pressure gradient (CFPG), but it has not been studied in complex CoA. Our objective was to characterize the effect of complex CoA and associated hemodynamics on CFPG in a clinical cohort.

**Methods and Results:** Retrospective analysis identified discrete juxtaductal (n=25) and complex CoA (n=43; transverse arch and/or isthmus hypoplasia) patients with arm-leg systolic blood pressure gradients (BPG) within 24 hours of echocardiography for comparison to BPG by conventional Doppler indices (simplified Bernoulli equation and modified forms correcting for proximal kinetic energy and/or recovered pressure). Results were interpreted using the current CoA guideline (BPG ≥20 mmHg) to compare diagnostic performance indicators including receiver operating characteristic curves, sensitivity, specificity, and diagnostic accuracy, among others. Echocardiography Z-scored aortic diameters were applied with computational stimulations from a preclinical CoA model to understand aspects of the CFPG driving performance differences.

Diagnostic performance was substantially reduced from discrete to complex CoA for conventional Doppler indices calculated from patient data, and by hypoplasia and/or long segment stenosis in simulations. In contrast, diagnostic indicators for the CFPG only modestly dropped for complex vs discrete CoA. Simulations revealed differences in performance due to inclusion of the Doppler velocity index and diastolic pressure half-time in the CFPG calculation.

**Conclusion:** CFPG is less affected by aortic arch anomalies co-existing with CoA when compared to conventional Doppler indices.

## Introduction

Coarctation of the aorta (CoA) is a constriction of the thoracic aorta and is one of the most common congenital cardiovascular defects^1–3^. While surgery is the optimal treatment in most infants, 5-30% of treated patients develop re-coarctation requiring reintervention^4^ and > 90% develop hypertension after 50 years of age^5^. Guidelines for intervention include a transcatheter peak-to-peak CoA gradient (BPGpp) ≥20 mmHg that is often regarded as hemodynamically significant^6^.

Despite allowing for direct measurement of the gradient, catheterization is an invasive procedure with potentially unwanted risks^7^. BPGpp can also be estimated via cuffs placed on the arms and legs. However, this is difficult in patients clinically due to children who may be upset, crying or scared during measurement, and/or inappropriately sized cuffs^8–10^. Moreover, CoA patients frequently have anomalies in their arterial anatomy that limit accurate determination of BPGpp via BP cuffs. For example, the left subclavian artery is hypoplastic in many patients, and branches distal to the coarctation in ∼5% of patients^11^. Non-invasive alternatives using continuous wave Doppler are therefore commonly applied in echocardiography to estimate the BPGpp^12^. This often involves the systolic gradient estimated from peak Doppler velocity using the simplified Bernoulli equation (SBE), or modified forms correcting for proximal kinetic energy (i.e., modified Bernoulli equation: MBE) and/or recovered pressure (SBE-RP)^13^. These conventional Doppler estimates are known to have poor agreement with catheterization^14^. Sources of disagreement include local discrepancy in aortic dynamic distensibility, simplification of the Bernoulli equation, and morphological variability, which can result in a substantial Doppler misclassification^1,15^. This misclassification is a critical barrier to assessing treatment modifications and translation since interpretation of new thresholds is best done using tools with a high degree of accuracy and diagnostic performance.

In contrast to tools exclusively focused on the systolic pressure gradient, we recently reported on a novel Doppler-based tool that accounts for the diastolic pressure gradient, which we termed the continuous flow pressure gradient (CFPG)^15^. Importantly, the CFPG was recently reported as an independent predictor of hemodynamic severity in the setting of discrete CoA devoid of complications found in complex CoA^1^. The physiologic underpinnings of a pronounced diastolic gradient can be explained by the prolonged recoil (i.e. capacitance) of the proximal aorta in response to the flow-limiting stenosis that leads to antegrade diastolic run-off with a typical sawtooth velocity decay^1^. As mentioned above, CoA in patients often includes hypoplasia in one or more arch segments presenting in concert with discrete CoA, or a long segment stenosis resulting in additional sites of local blood flow acceleration (i.e., complex CoA). The objective of the current study was to assess the diagnostic performance of the CFPG relative to conventional Doppler indices in a pediatric cohort of discrete and complex CoA cases. We further set out to determine the aspects of the CFPG that are driving differences in performance relative to conventional Doppler-based indices, which was conducted using Z-scored aortic diameters from the pediatric cohort in concert with representative computational hemodynamic simulations based on a preclinical model of CoA.

## Methods

### Study design and patient involvement

To identify the most relevant research topics and meaningful outcomes, the study was specially designed with the help of colleagues from the Division of Pediatric Cardiothoracic Surgery at Children’s Wisconsin and Herma Heart Institute, but no direct patient involvement was considered due to retrospective nature of the study design. After exempt determination by the IRB Board of Children’s Wisconsin, analysis of 259 CoA patients of either sex identified 25 cases with discrete juxtaductal CoA and 43 with complex CoA (transverse arch and/or isthmus hypoplasia), as well as contemporaneous arm-leg systolic blood pressure gradients via sphygmomanometry within 24 hours of echocardiography. Despite potential limitations^8–10^, sphygmomanometry was interpreted as a ground-truth for Doppler-based estimates of the gradient from CoA since catheter-based BP gradients were not available. Patients with patent ductus arteriosus, more than mild aortic valve insufficiency, other hemodynamically significant congenital heart diseases, low cardiac output state, and abnormal ventricular systolic function on echocardiography examination were excluded. Table 1 summarizes patient characteristics.

**Table 1-.**
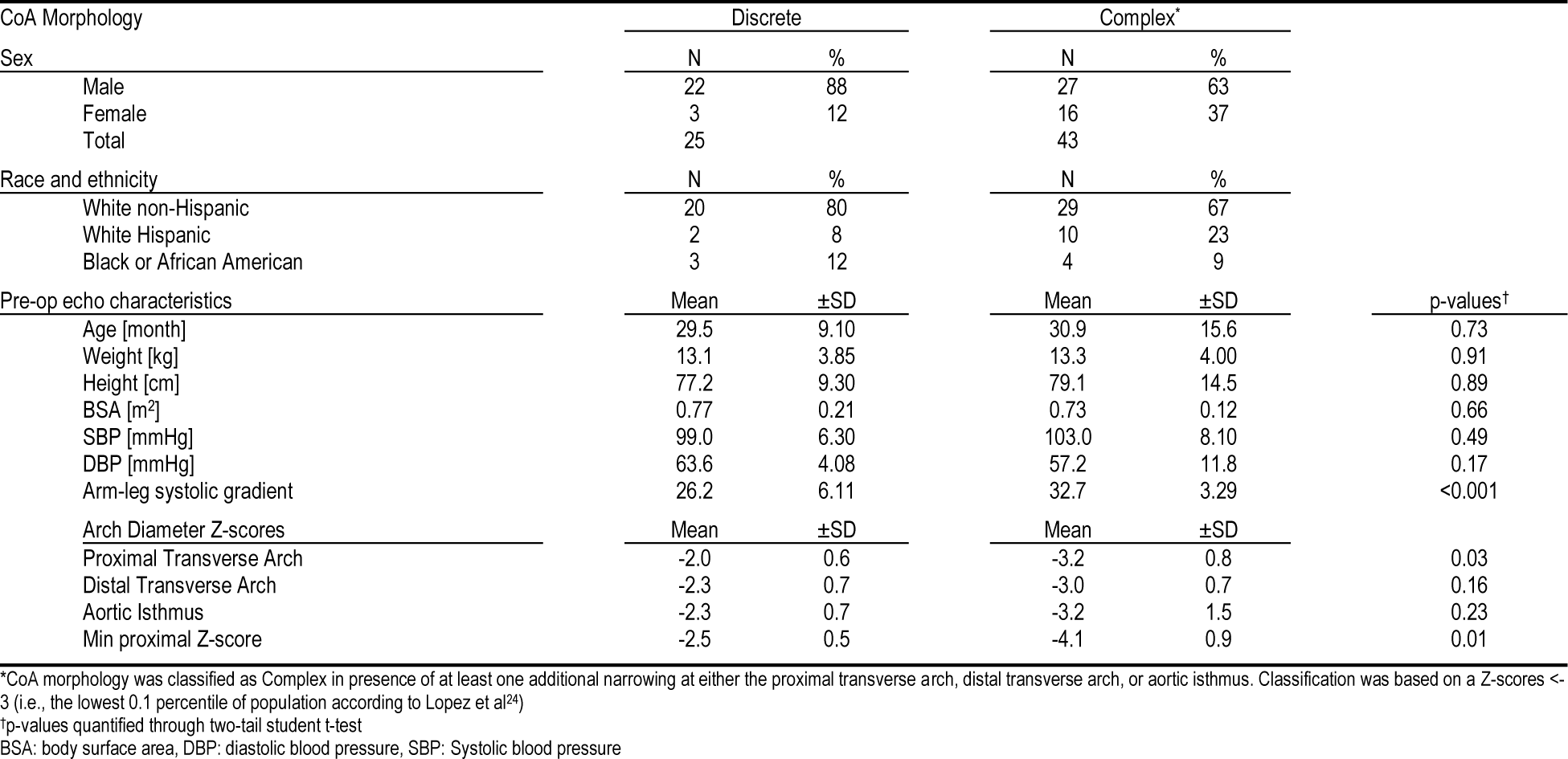
Characteristics for pediatric patients with discrete and complex CoA.

| CoA Morphology | Discrete |  | Complex* |  |  |
| --- | --- | --- | --- | --- | --- |
| Sex | N | % | N | % |  |
| Male | 22 | 88 | 27 | 63 |  |
| Female | 3 | 12 | 16 | 37 |  |
| Total | 25 |  | 43 |  |  |
| Race and ethnicity | N | % | N | % |  |
| White non-Hispanic | 20 | 80 | 29 | 67 |  |
| White Hispanic | 2 | 8 | 10 | 23 |  |
| Black or African American | 3 | 12 | 4 | 9 |  |
| Pre-op echo characteristics | Mean | ±SD | Mean | ±SD | p-values† |
| Age [month] | 29.5 | 9.10 | 30.9 | 15.6 | 0.73 |
| Weight [kg] | 13.1 | 3.85 | 13.3 | 4.00 | 0.91 |
| Height [cm] | 77.2 | 9.30 | 79.1 | 14.5 | 0.89 |
| BSA [m<sup>2</sup>] | 0.77 | 0.21 | 0.73 | 0.12 | 0.66 |
| SBP [mmHg] | 99.0 | 6.30 | 103.0 | 8.10 | 0.49 |
| DBP [mmHg] | 63.6 | 4.08 | 57.2 | 11.8 | 0.17 |
| Arm-leg systolic gradient | 26.2 | 6.11 | 32.7 | 3.29 | <0.001 |
| Arch Diameter Z-scores | Mean | ±SD | Mean | ±SD |  |
| Proximal Transverse Arch | -2.0 | 0.6 | -3.2 | 0.8 | 0.03 |
| Distal Transverse Arch | -2.3 | 0.7 | -3.0 | 0.7 | 0.16 |
| Aortic Isthmus | -2.3 | 0.7 | -3.2 | 1.5 | 0.23 |
| Min proximal Z-score | -2.5 | 0.5 | -4.1 | 0.9 | 0.01 |

\*CoA morphology was classified as Complex in presence of at least one additional narrowing at either the proximal transverse arch, distal transverse arch, or aortic isthmus. Classification was based on a Z-scores <-3 (i.e., the lowest 0.1 percentile of population according to Lopez et al<sup>24</sup>)
†p-values quantified through two-tail student t-test
BSA: body surface area, DBP: diastolic blood pressure, SBP: Systolic blood pressure

Echocardiographic measurements included conventional peak instantaneous pressure gradient estimation by SBE, MBE and SBE-RP^16^. An estimate of the pressure gradient during diastole was also calculated using diastolic flow continuation as characterized by the CFPG discussed in detail elsewhere^1^. All measurements were quantified from 2D and spectral Doppler images with early diastolic velocity identified at the end of the T wave^1^. Details related to echo quantification methods are summarized in Supplemental Table 1.

### Computational assessment of Doppler-derived index performance

Computational models were constructed using a pre-clinical rabbit model mimicking CoA in patients^17,18^ since measurements necessary to replicate physiology and aortic morphology beyond the coarctation region were not available in the echocardiography data from pediatric patients retrospectively studied. This approach allowed for understanding of hemodynamic factors impacting and/or potentially limiting the utility of the Doppler-based indices studied, including CFPG. Arterial segments were generated in SimVascular (simtk.org) using a control rabbit MRI dataset obtained following Institutional Animal Care and Use Committee approval^2^. The aortic diameter was then quantified at various locations throughout the thoracic aorta. This step was important to identify the normal population sized as baseline and realistically replicate discrete and complex CoA characteristics by matching Z-scores from computational models to the clinical datasets.

Model construction involved centerline detection, segmentation using a semi-automated level set method^19^, and lofting to create the blood flow domain. An in-house code was then developed to generate local modifications with the guidance of a pediatric cardiovascular surgeon by smoothly scaling aortic cross-sectional area according to shape functions^20^ representing different types of complex CoA including long-segment CoA, arch and isthmus hypoplasia (Supplemental Figure 1). The resulting morphological properties replicated Z-scores observed at the equivalent locations for each of the pediatric CoA patients^21^. A total of 68 computational aortas (25 with discrete CoA and 43 with complex CoA) were created. The aorta models were then integrated with arch branches (Supplemental Figure 2) for hemodynamic simulations with one inlet at the Sino-tubular junction and outlets at each of the head and neck arteries.

One-dimensional computational reduced order models (ROM) were generated in SimVascular (simvascular.github.io/) from the 3D CoA models^22^. The ROM approach was chosen for fast determination of hemodynamic results used to calculate Doppler indices (Supplemental Table 1) and automating the simulation process as compared to a semi-automated computationally intensive process required for 3D simulations with additional spatiotemporal information not necessary for the purpose of calculating Doppler-derived indies. A 3-element Windkessel model was implemented at each outlet to realistically replicate downstream pressure-flow relationships^23^ based on empirical measurements of pressure and flow from rabbits with discrete CoA. Windkessel model parameters were then adjusted to represent hemodynamics for complex CoA cases (see details in Supplemental Table 2). Data from discrete CoA rabbits with 42-96% area obstruction were used to replicate clinical BPGpp^18^ and the influence of CoA on thickness and stiffness after remolding of the wall that were included in computational simulations (see supplementary materials).

### Statistical analysis

Hypoplasia proximal to the CoA was quantitatively characterized using Z-score criteria^24^. Arch and isthmus hypoplasia was considered significant for Z-scores <-3.0 (i.e., dimensions in the lowest 0.1% of the population). Pearson’s correlation and linear regression were used to characterize the relationship between Doppler-based estimates and simulated pressure gradients. Adjusted R-squared values quantified the degree of scatter for each CoA severity and morphology studied. Diagnostic performances were compared via receiver operative characteristic (ROC) curves and relative to the current intervention guideline^17,25^ (i.e., BPGpp ≥20 mmHg). The optimum cutoffs for Doppler indices were identified by maximizing the sum of sensitivity and specificity, (i.e.; the Youden index^26^) and the area under the ROC curves (AUC) was compared using two-tailed student’s t-test with 5 and 10% significance levels. Finally, sensitivity, specificity, positive predictive values (PPV), negative predictive value (NPV), diagnostic odds ratio, and diagnostic accuracy were also quantified as measures of diagnostic performance. Reliability of the echo-based measurements used in calculating metrics including, SBE, MBE, SBE-RP, and indices used in the calculation of the CFPG including the diastolic pressure half-time (dPHT) and Doppler velocity index (DVI) was investigated using intraclass correlation coefficient (ICC) through SPSS software. Two observers were trained to quantify echo-based indices on a random subgroup of simulation and pediatric CoA patient datasets (n=10). Each metric was quantified in triplicate for inter-observer variability with median values used for intra-observer variability analysis^27^.

## Results

Doppler quantification from the cohort of pediatric CoA patients showed decreases in diagnostic performance moving from discrete to complex CoA morphologies, where correlation to proximal acceleration was evident. For example, the diagnostic accuracy was 0.88, 0.87, 0.79 for SBE, MBE, and SBE-RP indices from discrete CoA cases. These values dropped to 0.69, 0.59, and 0.59 for complex CoA cases, respectively. The diagnostic accuracy of CFPG was 0.96 for discrete CoA morphologies, and modestly dropped to 0.88 for complex CoA data sets. A detailed list of diagnostic performance indicators is provided in Table 2.

**Table 2-.**
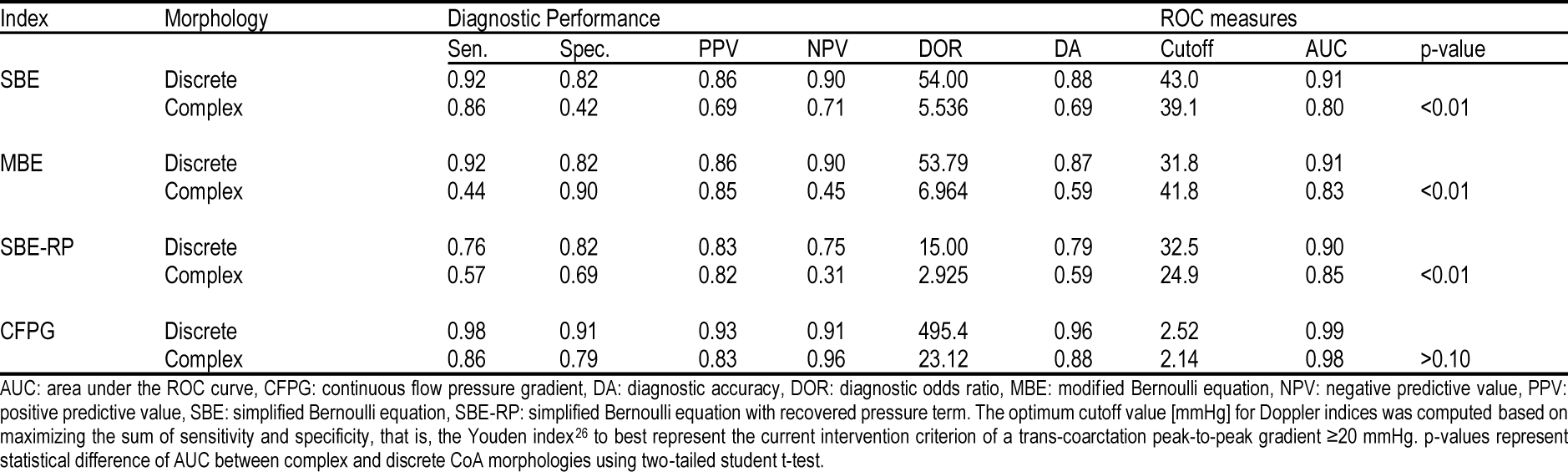
Diagnostic performance of Doppler-derived indices in pediatric CoA patients at optimum ROC cutoffs.

Similar patterns of reduced diagnostic performance were observed from the ROC analysis and AUCs. Specifically, the AUC was 0.91, 0.91, and 0.90 for conventional Doppler indices (i.e., SBE, MBE, and SBE-RP), which all significantly dropped to 0.80, 0.83, and 0.85, respectively (p-values <0.01, Table 2). On the other hand, the AUC for CFPG remained high at 0.98-0.99 despite additionally narrowing proximal to the coarctation for complex CoA cases (Table 2).

Reliability of the echo-based measurements used in calculating CFPG as quantified by ICC index revealed ≤9.8 and 11.2% differences among inter and intra-observer measurements, respectively. ICC index also confirmed good to excellent inter and intra-observer agreement in the conventional Doppler indices quantified. The 95% confidence interval for ICC was [0.86,0.98] for single subject variability and [0.91,0.99] on average, which is referred as excellent reliability according to Cicchetti et al^28^ and good according to Koo et al^27^.

Before assessing the diagnostic performance of each Doppler-based index quantified using echo data from the retrospective cohort of pediatric CoA patients via simulation, we first confirmed morphological similarity via Z-scores. Z-scores at the proximal transverse arch, distal transverse arch and aortic isthmus region were −3.22 ±0.81, −3.01 ±0.72, and −3.23 ±1.50 in the cohort of pediatric CoA patients, and corresponding values from computational models were −3.18 ±1.18, 3.11 ±0.52, and 3.17 ±1.68, which were not significantly different (p-values >0.1).

When compared to a discrete CoA, predictive values from conventional Doppler indices from ROM simulations were substantially reduced by the addition of hypoplasia and/or long segment stenosis in the arch. Specifically, the diagnostic accuracy was 0.63, 0.82, 0.80 for SBE, MBE, and SBE-RP indices for simulations with discrete CoA configurations. These values dropped to 0.48, 0.80, and 0.73 for simulations of complex CoA models, respectively. In contrast, the diagnostic accuracy of CFPG was 0.90 for simulations of discrete CoA morphologies, and only modestly dropped to 0.83 for simulations of complex CoA. A detailed list of predictive values is provided in Table 3.

**Table 3-.**
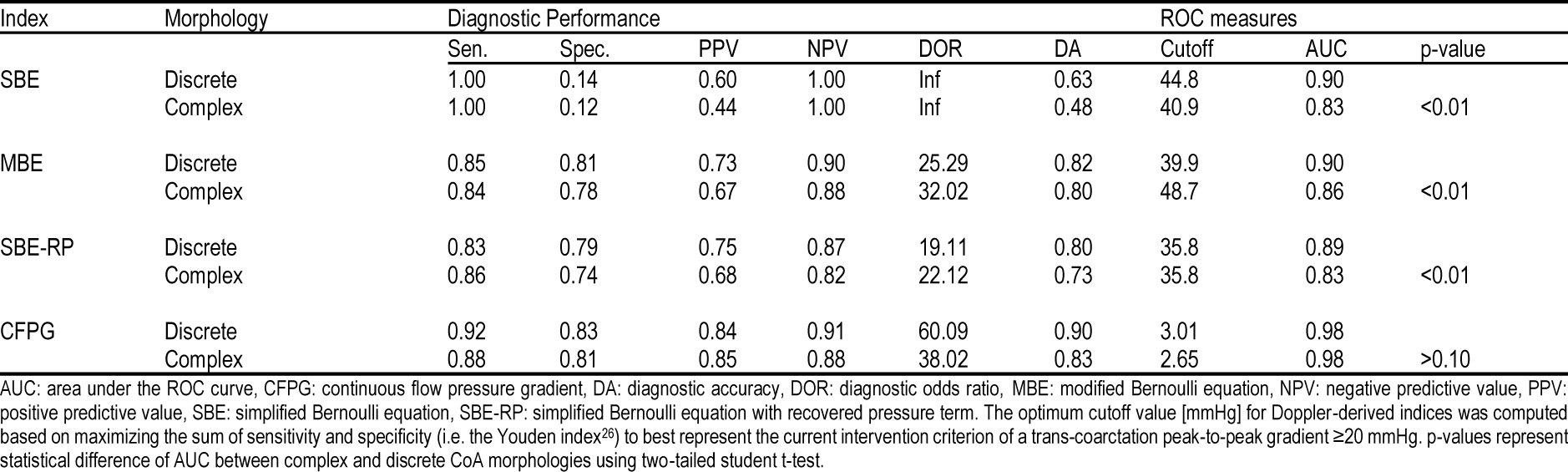
Diagnostic performance of Doppler-derived indices calculated at optimal ROC-derived cutoffs based on simulation results. Note the behavior from simulations is similar to data from pediatric CoA patients, with CFPG being a more consistent measure of diagnostic performance and ROC measures. This similarity allows for further analysis of those aspects of hemodynamics and performance impacting Doppler-based indices using the simulation results.

A similar pattern of reduced diagnostic performance was observed in AUCs calculated from ROC curves of Doppler indices determined by discrete CoA simulations (Figure 1, Table 3). Specifically, the AUC was 0.90, 0.90, and 0.89 for conventional Doppler indices (i.e., SBE, MBE, and SBE-RP) calculated from simulations, which significantly dropped to 0.83, 0.86, and 0.83, respectively (p-values <0.01, Table 2). Moving from discrete to complex CoAs, ROC analysis showed a pronounced shift towards non-diagnostic line (identity line, bottom left-to upper-right) for simulations of conventional Doppler indices. On the other hand, ROC curves obtained by CFPG calculated from simulations were relatively unchanged.

**Figure 1-.**
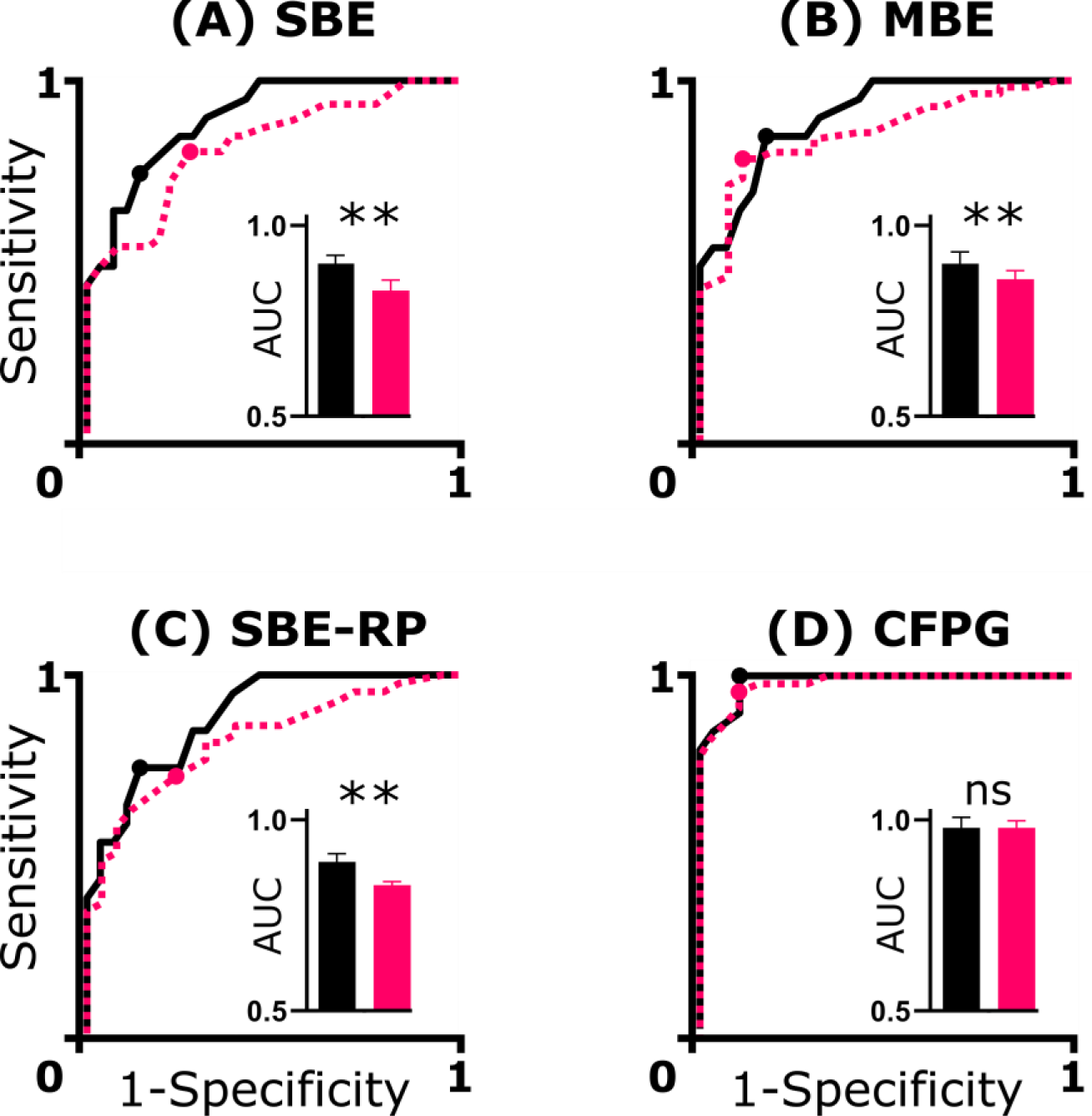
Receiver operative characteristic (ROC) curves for Doppler-derived indices of CoA severity calculated from simulations. CoA were classified according to the current clinical threshold (i.e., peak-to-peak trans-coarctation gradient ≥20 mmHg). The optimal cutoff value for Doppler indices, as reported in Table 3, was computed based on maximizing the sum of sensitivity and specificity (i.e. the Youden index^26^ indicated in dots for each ROC curve). Results are presented for Discrete (black-solid), and complex CoA morphologies (pink-dashed). CFPG: continuous flow pressure gradient, MBE: modified Bernoulli equation, RP: recovered pressure, SBE: simplified Bernoulli equation. Bar plots represent statistical difference of AUC between complex (pink) and discrete (black) CoA morphologies using student t-test with ** and ns indicating two-tail p-value <0.01 and >0.1.

For simulations of discrete CoA, regression analysis resulted in R^2^ = 0.88 for CFPG and 0.74-0.76 for conventional Doppler indices (Figure 2). Simulations of complex CoAs revealed the impact concomitant anomalies had on resulting flow patterns. For example, hemodynamic alterations including proximal acceleration and increased jet velocity were observed resulting in a higher degree of scatter (Figure 2). These factors decreased R^2^ values for conventional Doppler indices from simulations to 0.51-0.62, but to a lesser extent for CFPG calculated from simulations (0.78), indicating that CFPG outperformed conventional Doppler indices for simulations of both discrete and complex CoA.

**Figure 2-.**
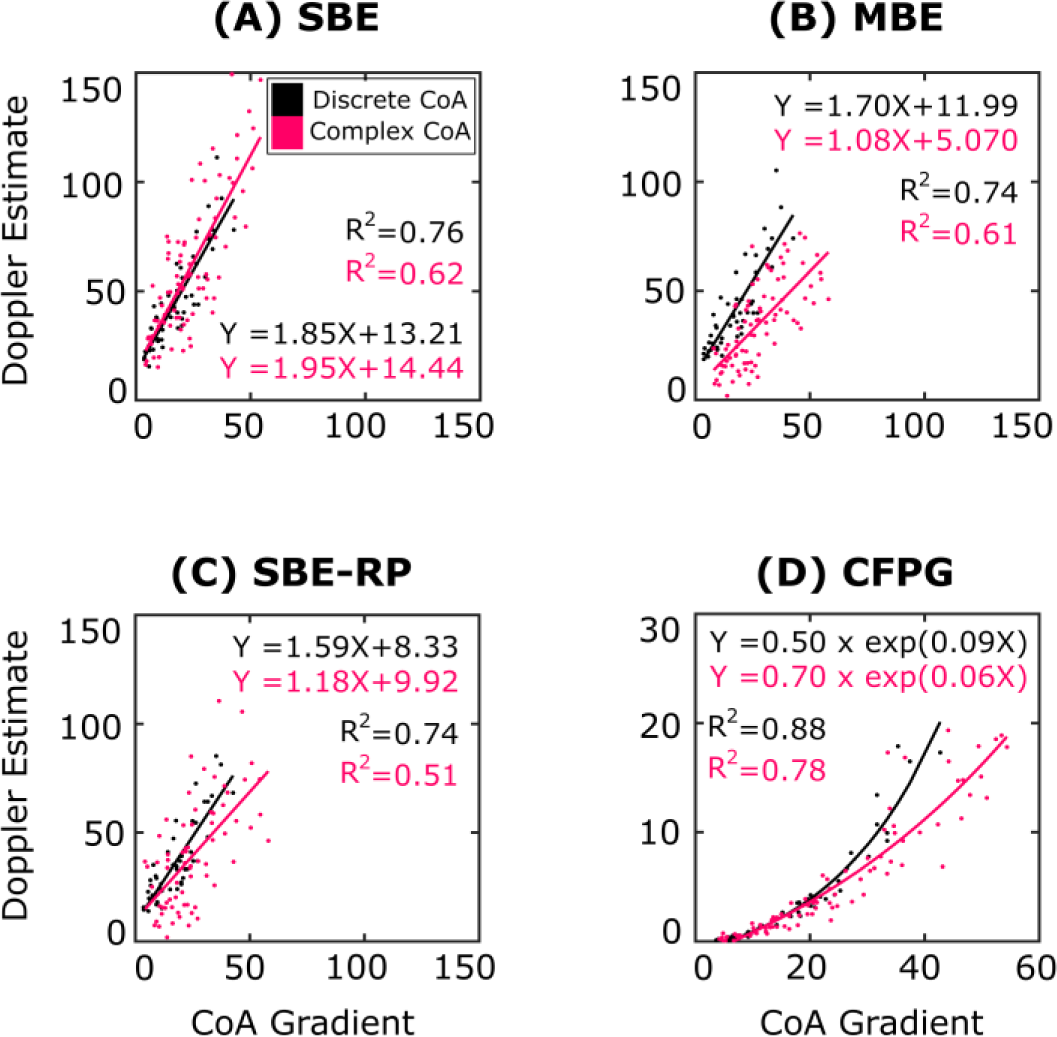
Comparison of Doppler-based indices from simulation results of discrete (black) and complex (pink) CoA morphologies. Doppler-derived indices (vertical-axis) were quantified from simulations using equations in Supplemental Table 1 including the (A) simplified Bernoulli equation (SBE) and its modified versions, i.e., (B) MBE, and (C) SBE-RP, as well as the continuous flow pressure gradient (CFPG). Adjusted R^2^ in linear regression analysis (solid lines) quantified degree of scatter resulting from each Doppler-derived index for discrete and complex CoA morphologies. The carotid artery to descending aorta pressure gradient was also quantified from simulation results to mimic the gold-standard CoA gradient from catheterization (horizontal axis). Differences in performance for Doppler indices are rooted in the fact that the CFPG includes several additional sub-indices described in Supplemental Table 1, namely the DVI and dPHT, which consider the hemodynamic changes resulting in complex CoA cases leading to a nonlinear correlation (Panel D and Supplemental Figure 3) more sensitive to higher CoA severities.

## Discussion

Suboptimal agreement with gold-standard catheterization represents a major limitation of conventional Doppler-derived indices used for non-invasive CoA severity assessment^14,29^. Although diastolic continuation of flow and estimation of its associated pressure gradient using the CFPG index previously showed strong correlation with measured values in a group of 25 discrete CoA patients^1^, its performance relative to conventional Doppler-derived indices had not been assessed in complex CoA morphologies. The main finding of the current study is that the CFPG was more consistent than any of the other Doppler-derived indices studied in the setting of complex CoA in a cohort of pediatric patients. It is also important to note that our results indicate the CFPG better correlates with arm-leg pressure gradients than the other Doppler-derived indices studied.

Performance differences among our novel CFPG method and conventional Doppler-derived indices can be understood relative to the contribution of local stenoses beyond discrete CoA using the simulations recounted above where physiologic boundary conditions were used to computationally investigate the diagnostic performance of CFPG and three conventional Doppler indices (SBE, MBE, and SBE-RP). Special care was taken to replicate morphological anomalies seen in complex forms of pediatric CoA patients by implementing similar Z-scores in proximal transverse arch, distal transverse arch, and aortic isthmus regions characterizing arch hypoplasia relative to the normal population^24^ (Table 1).

Differences in performance made clear by these simulations are rooted in the fact that the CFPG includes several additional sub-indices described in Supplemental Table 1, namely the DVI and dPHT, which consider the hemodynamic changes resulting in complex CoA cases. For example, in complex CoA, the DVI (proximal to jet peak velocity ratio) can increase due to proximal acceleration from a hypoplastic arch or isthmus (Figure 3, left). This leads to underestimation of CoA (Figure 2, B and C), and leads to discrepancy between discrete and complex CoA cases. However, the dPHT, which represents the time to half of the early diastolic pressure (*V_d_* to 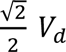 interval on the spectral doppler image, see Supplemental Table 1) accounts for the prolonged recoil of the proximal aorta and associated antegrade diastolic flow. This leads to consistent behavior of the dPHT in the setting of additional proximal stenoses in the form of a hypoplastic arch and/or isthmus (Figure 3, right).

**Figure 3-.**
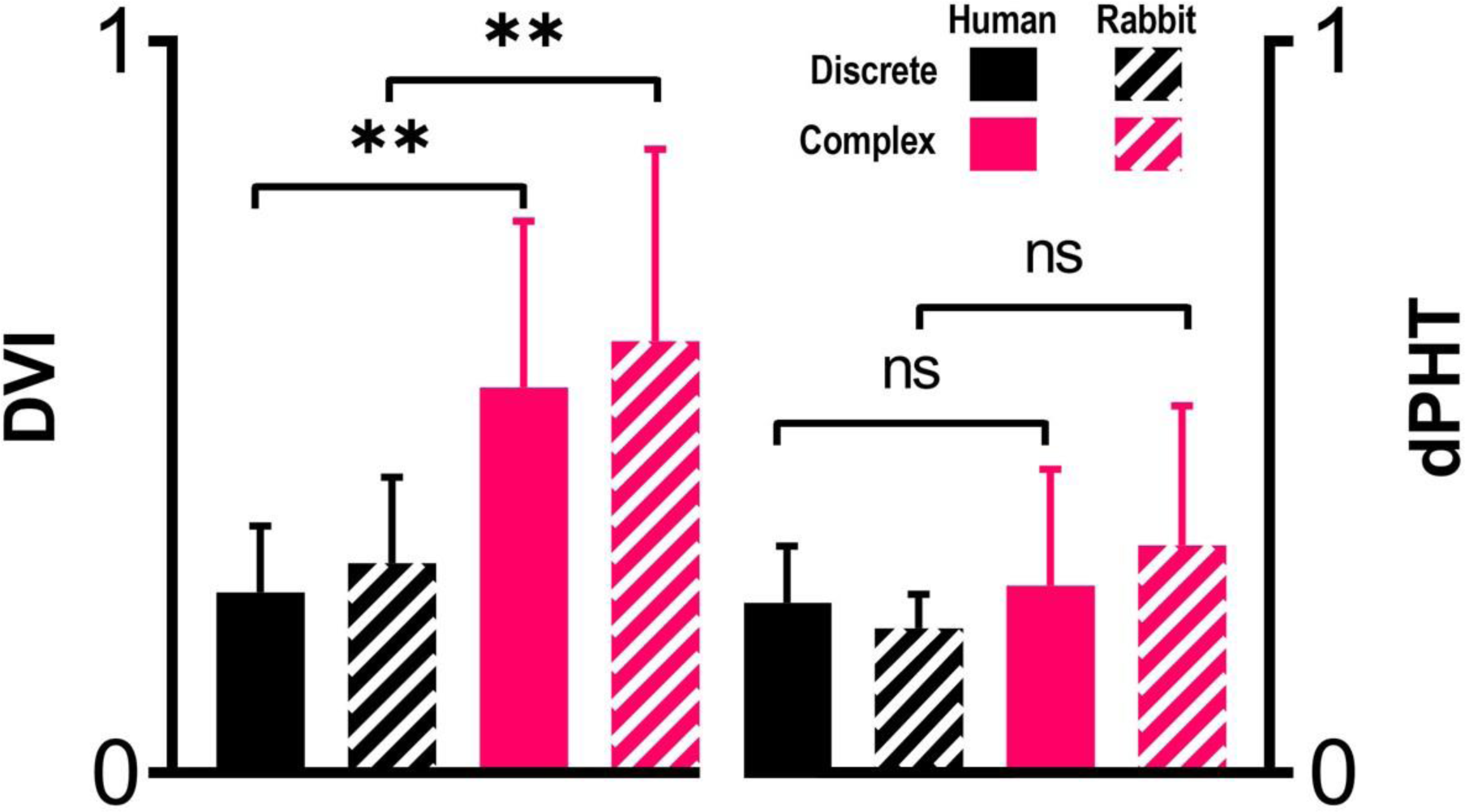
DVI (left) and dPHT (right) ratios for discrete (black) and complex (pink) CoA morphologies. No significant changes were observed in any of the indices between our cohort of pediatric CoA patients and simulations matching Z-scores for aortic morphology (p-value >0.1). There was a significant increase in DVI from discrete to complex CoA morphology in both our cohort of pediatric CoA patients (p-values <0.01), and simulations helped to reveal this finding is due to flow acceleration proximal to the CoA caused by hypoplastic arch, isthmus, or both. No significant differences were observed in dPHT among morphologies (p-values >0.1). p-values quantified using a two-tail student t-tests with 0.01 significance level.

Consequently, although a mixed behavior is possible due to the DVI and dPHT having opposite effects on the CFPG as indicated in Supplemental Figure 3, the CFPG is still relatively insensitive to proximal acceleration in the clinical ranges of complex CoA studied. Specifically, CFPG increases with dPHT making it more sensitive to additional proximal narrowing while, on the other hand, it decreases with DVI as is also seen in MBE and SBE-RP. Therefore, collectively the CFPG performance tends to be less affected by the proximal acceleration as suggested in the AUCs shown in Figure 1 and values reported in Table 3.

Pronounced overestimation of the peak instantaneous pressure gradient commonly reported in the literature for conventional Doppler-derived indices is further confirmed in the current work, leading to superficial sensitivity of 1.00 for SBE compromising specificity. Conversely, sensitivity values <0.14 will lead to a superficial diagnostic odds ratio (DOR) of infinity that positively correlates with balanced sensitivity and specificity under normal circumstance. This imbalance between sensitivity and specificity is particularly challenging in translational studies were intervention guidelines (e.g., CoA gradient) are ideally identified using invasive high-fidelity pressure measurements techniques such as catheterization, but results are being used in clinical setting based on non-invasive estimations such as echocardiography. Hence, the improved sensitivity-specificity balance observed in CFPG (Table 3) is promising evidence that this index can be of more potential value translationally. Particularly, in the setting complex CoA morphology, CFPG continues to deliver excellent balance between high sensitivity and specificity that is also evident in DORs when compared to conventional Doppler indices. In clinical practice, CFPG can be interpreted relative to the arm-leg cuff gradient identified using the current dataset. Table 4 summarized CFPG cutoffs for a range of clinically important CoA gradients.

**Table 4-.**
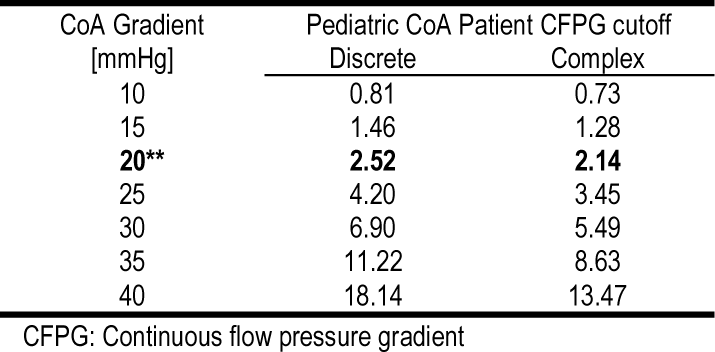
CFPG cutoffs for a range of clinically important CoA gradients.

The current results should be interpreted relative to several limitations. For example, literature suggests the arm-leg gradient may not be a strong predictor of invasive gradient in CoA patients^1,30^. Therefore, we expect the CFPG cutoffs obtained here may be different if sphygmomanometry data are compared to gold-standard catheterization data, which were not available for the cohort of pediatric CoA patients studied. Additionally, the cohort studied represented pediatric patients. Hence, it remains to be determined if the relationships presented in Tables 2 and 4 are also generalizable to older CoA patients. Complex morphologies were generated in computational representations of rabbit aortas by replicating Z-scores seen clinically at the thoracis aorta for CoA patients. This is in contrast to using measurements from the echo data within the CoA region of pediatric patients that would need to be coupled with assumptions related to blood flow distributions and dimensions of the ascending aorta and its branches. Considering these limitations, clinical validation of CFPG in preventing postsurgical complications including hypertension has yet to be studied in future work.

## Data Availability

All data produced in the present study are available upon reasonable request to the authors.

## Abbreviations

AA: Ascending aorta
BP: Blood pressure
CFPG: Continuous flow pressure gradient
CoA: Coarctation of the aorta
DBP: Diastolic blood pressure
DVI: Doppler velocity index
MBE: Modified Bernoulli equation
PC-MRI: Phase contrast magnetic resonance image
ROM: Reduced order model
SBE: Simplified Bernoulli equation
SBE-RP: SBE modified for recovered pressure
SBP: Systolic blood pressure

## Supplementary materials

Supplementary materials are available online.

## Acknowledgements

The authors gratefully acknowledge the technical support from Leanne Harman, Lindsey Kalvin, Jamasp Azarnoosh, and Chalani Ellepola.

## References

1. Ghorbannia A, Ellepola CD, Woods RK, Ibrahim E-SH, Maadooliat M, Ramirez HM, LaDisa JFJ. Clinical, Experimental, and Computational Validation of a New Doppler-Based Index for Coarctation Severity Assessment. J Am Soc Echocardiogr Off Publ Am Soc Echocardiogr 2022;35:1311–1321.

2. Azarnoosh J, Ghorbannia A, Ibrahim E-SH, Jurkiewicz H, Kalvin L, LaDisa JF. Temporal evolution of mechanical stimuli from vascular remodeling in response to the severity and duration of aortic coarctation in a preclinical model. Sci Rep 2023;13:8352.

3. Ghorbannia A, Maadooliat M, Woods RK, Audi SH, Tefft BJ, Chiastra C, Ibrahim ESH, LaDisa JF. Aortic Remodeling Kinetics in Response to Coarctation-Induced Mechanical Perturbations. Biomedicines 2023;11:1817.

4. Sweeney MS, Walker WE, Duncan JM, Hallman GL, Livesay JJ, Cooley DA. Reoperation for aortic coarctation: techniques, results, and indications for various approaches. Ann Thorac Surg 1985;40:46–49.

5. Kenny D, Polson JW, Martin RP, Paton JF, Wolf AR. Hypertension and coarctation of the aorta: an inevitable consequence of developmental pathophysiology. Hypertens Res 2011;34:543–547.

6. Feltes TF, Bacha E, Beekman RH, Cheatham JP, Feinstein JA, Gomes AS, Hijazi ZM, Ing FF, Moor M de, Morrow WR, Mullins CE, Taubert KA, Zahn EM. Indications for Cardiac Catheterization and Intervention in Pediatric Cardiac Disease. Circulation 2011;123:2607–2652.

7. Yang JC-T, Lin M-T, Jaw F-S, Chen S-J, Wang J-K, Shih TT-F, Wu M-H, Li Y-W. Trends in the utilization of computed tomography and cardiac catheterization among children with congenital heart disease. J Formos Med Assoc Taiwan Yi Zhi 2015;114:1061–1068.

8. Álvarez J, Aguilar F, Lurbe E. Blood pressure measurement in children and adolescents: key element in the evaluation of arterial hypertension. An Pediatr 2022;96:536.e1–536.e7.

9. Bird C, Michie C. Measuring blood pressure in children. BMJ 2008;336:1321.

10. Palatini P, Asmar R. Cuff challenges in blood pressure measurement. J Clin Hypertens Greenwich Conn 2018;20:1100–1103.

11. Agasthi P, Pujari SH, Tseng A, Graziano JN, Marcotte F, Majdalany D, Mookadam F, Hagler DJ, Arsanjani R. Management of adults with coarctation of aorta. World J Cardiol 2020;12:167–191.

12. Lim DS, Ralston MA. Echocardiographic indices of Doppler flow patterns compared with MRI or angiographic measurements to detect significant coarctation of the aorta. Echocardiogr Mt Kisco N 2002;19:55–60.

13. Giardini A, Tacy TA. Pressure recovery explains doppler overestimation of invasive pressure gradient across segmental vascular stenosis. Echocardiography 2010;27:21–31.

14. Marx GR, Allen HD. Accuracy and pitfalls of Doppler evaluation of the pressure gradient in aortic coarctation. J Am Coll Cardiol 1986;7:1379–1385.

15. Ghorbannia A, Ellepola CD, Woods RK, LaDisa JF. Abstract 10882: Validation of a Continuous Flow Pressure Gradient for Coarctation Severity Assessment. Circulation 2021;144:A10882–A10882.

16. Singh GK, Mowers KL, Marino C, Balzer D, Rao PS. Effect of Pressure Recovery on Pressure Gradients in Congenital Stenotic Outflow Lesions in Pediatric Patients-Clinical Implications of Lesion Severity and Geometry: A Simultaneous Doppler Echocardiography and Cardiac Catheter Correlative Study. J Am Soc Echocardiogr Off Publ Am Soc Echocardiogr 2020;33:207–217.

17. Menon A, Eddinger TJ, Wang H, Wendell DC, Toth JM, LaDisa Jr. JF. Altered hemodynamics, endothelial function, and protein expression occur with aortic coarctation and persist after repair. Am J Physiol Heart Circ Physiol 2012;303:H1304–18.

18. Menon A, Wendell DC, Wang H, Eddinger TJ, Toth JM, Dholakia RJ, Larsen PM, Jensen ES, Ladisa Jr. JF. A coupled experimental and computational approach to quantify deleterious hemodynamics, vascular alterations, and mechanisms of long-term morbidity in response to aortic coarctation. J Pharmacol Toxicol Methods 2012;65:18–28.

19. Updegrove A, Wilson NM, Merkow J, Lan H, Marsden AL, Shadden SC. SimVascular: An Open Source Pipeline for Cardiovascular Simulation. Ann Biomed Eng 2017;45:525–541.

20. Ghorbanniahassankiadeh A, Marks DS, LaDisa JF Jr. Correlation of Computational Instantaneous Wave-Free Ratio With Fractional Flow Reserve for Intermediate Multivessel Coronary Disease. J Biomech Eng 2021;143.

21. Teien DE, Wendel H, Björnebrink J, Ekelund L. Evaluation of anatomical obstruction by Doppler echocardiography and magnetic resonance imaging in patients with coarctation of the aorta. Br Heart J 1993;69:352–355.

22. Pfaller MR, Pham J, Verma A, Pegolotti L, Wilson NM, Parker DW, Yang W, Marsden AL. Automated generation of 0D and 1D reduced-order models of patient-specific blood flow. Int J Numer Methods Biomed Eng 2022;n/a:e3639.

23. LaDisa JFJ, Alberto Figueroa C, Vignon-Clementel IE, Kim HJ, Xiao N, Ellwein LM, Chan FP, Feinstein JA, Taylor CA. Computational simulations for aortic coarctation: representative results from a sampling of patients. J Biomech Eng 2011;133:091008.

24. Lopez L, Colan S, Stylianou M, Granger S, Trachtenberg F, Frommelt P, Pearson G, Camarda J, Cnota J, Cohen M, Dragulescu A, Frommelt M, Garuba O, Johnson T, Lai W, Mahgerefteh J, Pignatelli R, Prakash A, Sachdeva R, Soriano B, Soslow J, Spurney C, Srivastava S, Taylor C, Thankavel P, Velde M van der, Minich L. Relationship of Echocardiographic Z Scores Adjusted for Body Surface Area to Age, Sex, Race, and Ethnicity. Circ Cardiovasc Imaging 2017;10:e006979.

25. A. WC, G. WR, M. BT, S. CJ, M. CH, A. DJ, Pedro del N, W. FJ, P. GT, M. HZ, A. HS, Etta KM, J. LM, D. MP, J. RM, P. WE, D. WG. ACC/AHA 2008 Guidelines for the Management of Adults With Congenital Heart Disease. Circulation 2008;118:e714–e833.

26. Youden WJ. Index for rating diagnostic tests. Cancer 1950;3:32–35.

27. Koo TK, Li MY. A Guideline of Selecting and Reporting Intraclass Correlation Coefficients for Reliability Research. J Chiropr Med 2016;15:155–163.

28. Cicchetti D V. Guidelines, criteria, and rules of thumb for evaluating normed and standardized assessment instruments in psychology. Psychol Assess 1994;6:284–290.

29. Vlahos AP, Marx GR, McElhinney D, Oneill S, Goudevenos I, Colan SD. Clinical utility of Doppler echocardiography in assessing aortic stenosis severity and predicting need for intervention in children. Pediatr Cardiol 2008;29:507–514.

30. Patankar N, Fernandes N, Kumar K, Manja V, Lakshminrusimha S. Does measurement of four-limb blood pressures at birth improve detection of aortic arch anomalies? J Perinatol Off J Calif Perinat Assoc 2016;36:376–380.

