## Supplemental material for "Consistency of the continuous flow pressure gradient despite aortic arch anomalies co-existing with coarctation"

<sup>5</sup>Departments of Physiology, and Medicine - Division of Cardiovascular Medicine, Medical College
of Wisconsin, Milwaukee, Wisconsin, USA

**Address for correspondence:** Arash Ghorbannia, 8701 W Watertown Plank Rd., Milwaukee, WI

*Supplemental Table 1 - Calculations applied for peak instantaneous pressure gradient estimation by Doppler indices.*

| Doppler Estimates of CoA Gradient | Equation |
| --- | --- |
| <u>Systolic CoA Gradient</u> |  |
| SBE | $4v_{\text{peak}}^2$ |
| MBE | $4v_{\text{peak}}^2 - 4v_{\text{prox}}^2$ |
| SBE-RP | $8v_{\text{peak}}^2 \times A(1 - A),$<br>$A = A_{\text{CoA}}/A_{\text{prox}}$ |
| <u>Diastolic CoA Gradient</u> |  |
| CFPG | $4B \times \left(1 - \exp\left(-\frac{1}{B}\right)\right)(1 - \text{DVI})^2 v_d^2,$<br>$B = \text{dPHT}/(\ln(2) \times \text{DP})$<br>$\text{DVI} = v_{\text{prox}}/v_{\text{peak}}$ |

CFPG: continuous flow pressure gradient, DP: diastolic period, dPHT: diastolic pressure half-time, DVI: Doppler velocity index, MBE: modified Bernoulli equation, RP: recovered pressure, SBE: simplifies Bernoulli equation, $v_{\text{peak}}$: peak jet velocity at the CoA region obtained from spectral Doppler image, $v_{\text{prox}}$: peak velocity obtained proximal to the CoA from spectral Doppler image, $A_{\text{CoA}}$: CoA cross-section area obtained from Doppler b-mode image, $A_{\text{prox}}$: aorta cross-section area proximal to the CoA obtained from Doppler b-mode image, $v_d$: early diastolic velocity<sup>‡</sup>

<sup>†</sup>End T-Q interval on the EKG signal

<sup>‡</sup>Time to 50% early diastolic pressure; on the spectral Doppler image, time from $v_d$ to $\frac{\sqrt{2}}{2}v_d$

<sup>§</sup>Velocity at the end of T-wave obtained from cardiac-gated spectral Doppler image

<u>**Boundary conditions.**</u> After Institutional Care and Use Committee approval, CoA was surgically induced in New Zealand white rabbits to replicate clinical peak-to-peak blood pressure gradients<sup>1</sup> and empirically assess its effect on thickness and stiffness via remodeling of the wall. Approximately 22 weeks later and prior to terminal tissue harvest, time-resolved 2D through-plane PC-MRI was performed orthogonal to the head and neck arteries and ascending aorta just distal to the aortic valve to calculate total aortic flow and distribution to major branches<sup>2</sup> for discrete CoA rabbits of varying severities (n = 55). Image-based eddy current compensation was then performed in Segment (Medviso AB, version 2.0 R5152) to account for magnetic field inhomogeneity and associated uncertainty in flow quantification<sup>3-5</sup>. The aorta was then harvested after humane euthanasia. Material properties and wall thickness were quantified at the proximal ascending, proximal descending, and distal descending thoracic aorta at the diaphragm level. Material characterization was performed using uniaxial extension testing (MTS Criterion Load Frame,

MTS, Minneapolis, U.S.A.) to obtain stress-strain curves used to identify vessel stiffness<sup>6</sup> while wall thickness was quantified through sample preparation process and optically under the dissection scope. Regression analysis identified the thickness and stiffness values used for each computational model at 40, 60, 70, 80, and 90% CoA area obstructions Supplemental Figure 1; Supplemental Table 2). An in-house python script was developed to spatially distribute the wall characteristics obtained at location 1-3 of Supplemental Figure 1 as a linear function of aortic centerline.

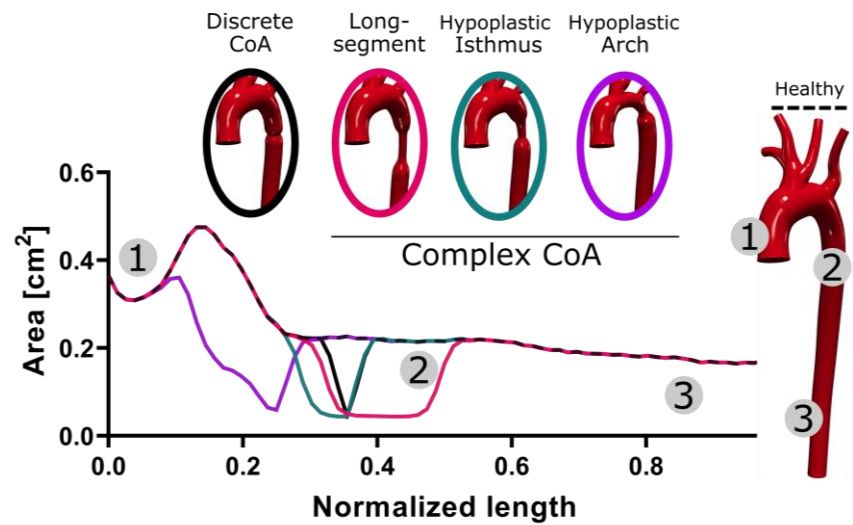

*Supplemental Figure 1- Longitudinal areas for 3D CAD models of discrete and complex CoA in rabbit aorta. Generalized models of CoA were created by smoothly scaling local cross-sectional area according to a sigmoid function of centerline length. Discrete (black) and complex CoA, i.e., long segment (pink), hypoplastic isthmus (green), and hypoplastic arch (purple), morphologies were created for the severities seen clinically. Locations 1, 2, and 3 correspond to the ascending, descending and distal thoracic aorta, respectively.*

Morphological properties for computational simulations replicated Z-scores observed at the equivalent locations for each of the pediatric CoA patients. A total of 68 computational aortas (25 with discrete CoA and 43 with complex CoA) were created. The aorta models were then integrated with arch branches (Supplemental Figure 2) for hemodynamic simulations with one inlet at the Sino-tubular junction and outlets at each of the head and neck arteries.

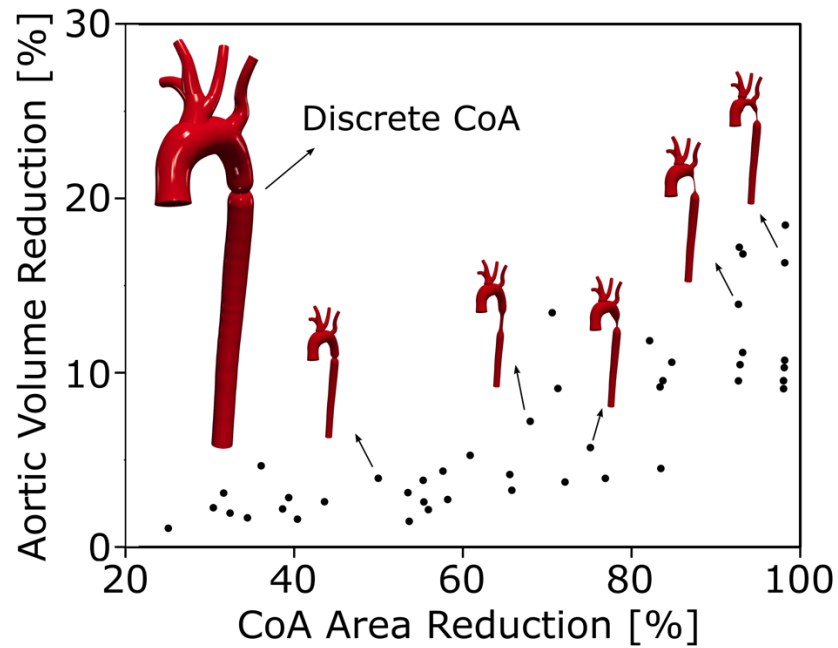

*Supplemental Figure 2- Three-dimensional CAD models for discrete and complex CoA morphologies generated from a rabbit aorta based on a clinical cohort of Z-scores in the arch and isthmus regions. Corresponding CoA models were created by smoothly scaling local cross-sectional area according to a sigmoid function of centerline length and corresponding aortic volume loss and CoA area reduction were normalized to that of control model for x and y axis visualization, respectively.*

High-fidelity blood pressure (BP) measurements were quantified from rabbit datasets with discrete CoA. BP waveforms were simultaneously measured proximal and distal to the CoA with the same model transducer (Harvard Apparatus, Holliston, MA) at 360 Hz using a computer interfaced with an analog-to-digital converter. Resulting waveforms were used to quantify mean arterial BP and associated time-constants<sup>7,8</sup>.

Three-element Windkessel models were used to replicate the impact of vessels downstream of aortic outlets using three parameters with physiologic meaning: characteristic resistance ($R_c$), capacitance ($C$), and distal resistance ($R_d$). The total systemic capacitance ($C_{total}$) was determined from the range of measured time constants<sup>8,9</sup> assuming a characteristic to total resistance ratio ($R_c/R_{total}$) of $\leq 11\%$<sup>9</sup>. Total systemic resistance ($R_{total} = R_c + R_d$) was identified from mean aortic flow and mean arterial pressure, i.e., $R_{total} = MAP/Q_{mean}$ (Supplemental Table 2). Linear regression characterized the correlation of total systemic resistance and capacitance with CoA severity. The physiological range of Windkessel parameters was then identified at each severity level (i.e., 40, 60, 70, 80, and 90%) from the 95% confidence interval of linear regression fits. Total systemic capacitance and resistance were then distributed to each outlet based on a generalization of Murray's law with ratios proportionate to the outlet cross-section areas<sup>10</sup>.

*Supplemental Table 2- Physiological range of boundary condition and vessel characteristics quantified in rabbits with CoA.*

| % Area Obstruction |  | 40 | 60 | 70 | 80 | 90 |
| --- | --- | --- | --- | --- | --- | --- |
| $R_{total}$ [$10^3 \times cgs$] | | [12.3,13.7] | [13.0,13.9] | [13.3,15.6] | [13.7,15.9] | [13.9,16.1] |
| Normalized time constant |  | [1.55,2.24] | [1.29,1.83] | [1.20,1.69] | [1.03,1.52] | [0.82,1.40] |
| $C_{total}$ [$10^{-5} \times cgs$] | Discrete | [3.79,5.43] | [3.25,4.34] | [2.82,3.96] | [2.33,3.57] | [1.82,3.11] |
|  | Complex | [3.79,4.14] | [3.25,3.69] | [2.82,3.37] | [2.33,2.91] | [1.82,2.45] |
| Wall Thickness [mm] | Location 1 | 0.48 ± 0.05 | 0.52 ± 0.03 | 0.54 ± 0.02 | 0.56 ± 0.03 | 0.59 ± 0.02 |
|  | Location 2 | 0.30 ± 0.03 | 0.30 ± 0.03 | 0.31 ± 0.03 | 0.34 ± 0.02 | 0.35 ± 0.02 |
|  | Location 3 | 0.31 ± 0.01 | 0.31 ± 0.01 | 0.26 ± 0.01 | 0.32 ± 0.01 | 0.30 ± 0.03 |
| Wall Stiffness [$10^6 \times cgs$] | Location 1 | 3.51 ± 0.23 | 7.03 ± 0.32 | 17.1 ± 0.21 | 21.5 ± 0.77 | 20.1 ± 0.25 |
|  | Location 2 | 7.53 ± 0.28 | 8.20 ± 0.32 | 16.9 ± 0.39 | 19.8 ± 0.81 | 18.6 ± 0.44 |
|  | Location 3 | 12.1 ± 0.37 | 15.2 ± 0.44 | 20.0 ± 0.72 | 22.2 ± 0.56 | 11.0 ± 0.33 |

*cgs: centimeter, gram, second. Locations 1, 2, and 3 indicate proximal ascending, proximal descending, and distal descending thoracic aorta, respectively. Data represent 95% confidence interval for total systemic resistance ($R_{total}$) and capacitance ($C_{total}$). Spatial distribution of wall thickness and stiffness are represented as mean ± S.D.*

Traditionally, tuning of computational arterial simulations is an iterative process starting from an initial guess for boundary conditions and with tweaking of parameters as needed, often manually, until resulting BP and flow distributions are within an acceptable range of target/measured values<sup>11–13</sup>. However, in the current study the physiological range of boundary conditions were known (Supplemental Table 2) from a group of rabbits with varying discrete CoA severity<sup>14–16</sup>. Therefore, a python script was developed to systematically evaluate possible combinations of boundary condition parameters by discretizing the solution space for total systemic resistance and capacitance as mentioned above. At each outlet the range of boundary condition parameters was discretized into five values, i.e., 5, 25, 50, 75, and 95<sup>th</sup> percentile. Similarly, the physiological range of $R_c/R_{total}$ was discretized to 0.1, 3, 6, 9, and 11%. Finally, to account for local coarctation-induced perturbation in the distribution of resistances and capacitances, -50, -25, 0, 25, and 50% of the total resistance and capacitances were redistributed to achieve values inferred from literature<sup>17–22</sup> as a result of the local constriction. This method created 5<sup>5</sup> possible choices for boundary condition parameters in the physiological range identified from all rabbit data referenced for the current study. One-dimensional ROM of blood flow were created for physiological boundary conditions and each complex CoA geometry. Results within 10% of the aimed pressure and flows for all outlets were used for post-processing and identification of Doppler-derived indices. Simulations were performed in SimVascular (simvascular.github.io/) using a discontinuous Galerkin space-time finite element method with piecewise linear shape functions<sup>23</sup>. Convergence criteria included residuals $<1\text{E-}4$. Each simulation was run for 6 cardiac cycles until results were periodic<sup>24</sup>. A total of 48 simulations met convergence criteria for discrete CoA morphology. For complex CoA, 28 simulations met the convergence criteria for long-segment and isthmus hypoplasia, and 27 for arch hypoplasia models.

For complex CoA cases, literature was used to identify a sub-range of parameter values from those identified for discrete CoA rabbits. More specifically, the extent of arterial stiffening, impaired distensibility, and changes in pulse pressure are expected to be more prominent for complex CoA cases<sup>17-22</sup>. This results in higher slopes for the pressure drop over the diastolic phase, increases the time-constant, and decreases the capacitance of the system<sup>7,8,25</sup>. Hence, a left-sided subregion of the 95% confidence intervals identified for discrete CoA was assumed to be representative of parameters for use with complex CoA simulations (see Complex in Supplemental Table 2).

Differences in performance for Doppler indices made clear by computational simulations are rooted in the fact that the CFPD includes several additional sub-indices described in Supplemental Table 1, namely the DVI and dPHT, which consider the hemodynamic changes resulting in complex CoA cases. Consequently, although a mixed behavior is possible due to the DVI and dPHT having opposite effects on the CFPD as indicated in Supplemental Figure 3, the CFPD is still relatively insensitive to proximal acceleration in the clinical ranges of complex CoA studied. Specifically, CFPG increases with dPHT making it more sensitive to additional proximal narrowing while, on the other hand, it decreases with DVI as is also seen in MBE and SBE-RP. Collectively, the CFPG performance tends to be less affected by the proximal acceleration.

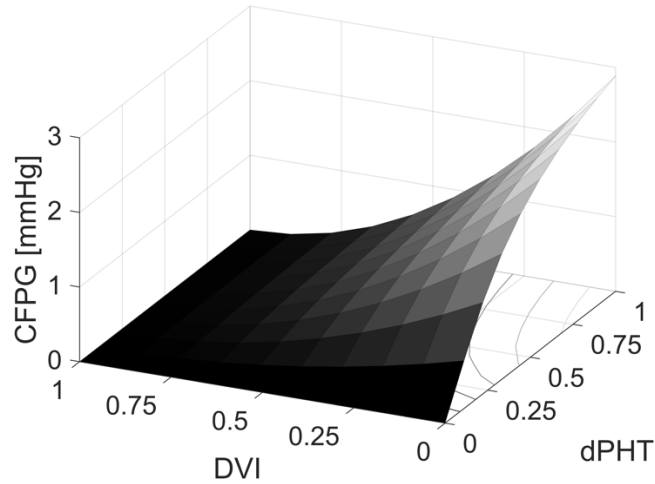

*Supplemental Figure 3- CFPG behavior vs. DVI and dPHT for a representative simulation case of 1 m/s early diastolic velocity. CFPG increases with dPHT making it more sensitive to additional proximal narrowing while it decreases with DVI as is also seen in MBE and SBE-RP. Therefore, collectively the performance of the CFPG tends to be less affected by the proximal acceleration seen in the forms of complex CoA studied.*

### References:

1. Menon A, Wendell DC, Wang H, et al. A coupled experimental and computational approach to quantify deleterious hemodynamics, vascular alterations, and mechanisms of long-term morbidity in response to aortic coarctation. *Journal of pharmacological and toxicological methods*. 2012;65(1):18-28. doi:10.1016/j.vascn.2011.10.003
2. Wendell DC, Friehs I, Samyn MM, Harmann LM, LaDisa JFJ. Treating a 20 mm Hg gradient alleviates myocardial hypertrophy in experimental aortic coarctation. *J Surg Res*. 2017;218:194-201. doi:10.1016/j.jss.2017.05.053
3. Lorenz R, Bock J, Snyder J, Korvink JG, Jung BA, Markl M. Influence of eddy current, Maxwell and gradient field corrections on 3D flow visualization of 3D CINE PC-MRI data. *Magn Reson Med*. 2014;72(1):33-40. doi:10.1002/mrm.24885
4. Bock J, Frydrychowicz A, Lorenz R, et al. In vivo noninvasive 4D pressure difference mapping in the human aorta: phantom comparison and application in healthy volunteers and patients. *Magn Reson Med*. 2011;66(4):1079-1088. doi:10.1002/mrm.22907
5. Zotti A, Banzato T, Cozzi B. Cross-sectional anatomy of the rabbit neck and trunk: comparison of computed tomography and cadaver anatomy. *Res Vet Sci*. 2009;87(2):171-176. doi:10.1016/j.rvsc.2009.02.003
6. Macrae RA, Miller K, Doyle BJ. Methods in Mechanical Testing of Arterial Tissue: A Review. *Strain*. 2016;52(5):380-399. doi:10.1111/str.12183
7. Chemla D, Lau EMT, Hervé P, et al. Influence of critical closing pressure on systemic vascular resistance and total arterial compliance: A clinical invasive study. *Archives of Cardiovascular Diseases*. 2017;110(12):659-666. doi:10.1016/j.acvd.2017.03.008
8. Kaiser AD, Shad R, Hiesinger W, Marsden AL. A design-based model of the aortic valve for fluid-structure interaction. *Biomech Model Mechanobiol*. 2021;20(6):2413-2435. doi:10.1007/s10237-021-01516-7
9. Laskey WK, Parker HG, Ferrari VA, Kussmaul WG, Noordergraaf A. Estimation of total systemic arterial compliance in humans. *J Appl Physiol (1985)*. 1990;69(1):112-119. doi:10.1152/jappl.1990.69.1.112
10. Zhou Y, Kassab GS, Molloy S. On the design of the coronary arterial tree: a generalization of Murray's law. *Phys Med Biol*. 1999;44(12):2929-2945. doi:10.1088/0031-9155/44/12/306
11. Menon A, Eddinger TJ, Wang H, Wendell DC, Toth JM, LaDisa Jr. JF. Altered hemodynamics, endothelial function, and protein expression occur with aortic coarctation and persist after repair. *American journal of physiology Heart and circulatory physiology*. 2012;303(11):H1304-18. doi:10.1152/ajpheart.00420.2012

12. LaDisa JFJ, Alberto Figueroa C, Vignon-Clementel IE, et al. Computational simulations for aortic coarctation: representative results from a sampling of patients. *J Biomech Eng.* 2011;133(9):091008. doi:10.1115/1.4004996
13. Kim HJ, Vignon-Clementel IE, Coogan JS, Figueroa CA, Jansen KE, Taylor CA. Patient-specific modeling of blood flow and pressure in human coronary arteries. *Ann Biomed Eng.* 2010;38(10):3195-3209. doi:10.1007/s10439-010-0083-6
14. Ghorbannia A, Maadooliat M, Woods RK, et al. Aortic Remodeling Kinetics in Response to Coarctation-Induced Mechanical Perturbations. *Biomedicines.* 2023;11(7):1817. doi:10.3390/biomedicines11071817
15. Azarnoosh J, Ghorbannia A, Ibrahim ESH, Jurkiewicz H, Calvin L, LaDisa JF. Temporal evolution of mechanical stimuli from vascular remodeling in response to the severity and duration of aortic coarctation in a preclinical model. *Scientific Reports.* 2023;13(1):8352. doi:10.1038/s41598-023-34400-8
16. Ghorbannia A, Ellepola CD, Woods RK, et al. Clinical, Experimental, and Computational Validation of a New Doppler-Based Index for Coarctation Severity Assessment. *J Am Soc Echocardiogr.* 2022;35(12):1311-1321. doi:10.1016/j.echo.2022.09.006
17. Ghorbani N, Muthurangu V, Khushnood A, et al. Impact of valve morphology, hypertension and age on aortic wall properties in patients with coarctation: a two-centre cross-sectional study. *BMJ open.* 2020;10(3):e034853. doi:10.1136/bmjopen-2019-034853
18. Wang J, Yang C, Lai B. Long-segmental middle aortic coarctation: a rare case first diagnosed by transthoracic echocardiography. *BMC Cardiovascular Disorders.* 2022;22(1):27. doi:10.1186/s12872-022-02475-2
19. Carvalho JS, Redington AN, Shinebourne EA, Rigby ML, Gibson D. Continuous wave Doppler echocardiography and coarctation of the aorta: gradients and flow patterns in the assessment of severity. *Heart.* 1990;64(2):133. doi:10.1136/hrt.64.2.133
20. Yu Y, Wang Y, Yang M, et al. Evaluating the severity of aortic coarctation in infants using anatomic features measured on CTA. *Eur Radiol.* 2021;31(3):1216-1226. doi:10.1007/s00330-020-07238-1
21. Hajsadeghi S, Fereshtehnejad SM, Ojaghi M, et al. Doppler echocardiographic indices in aortic coarctation: a comparison of profiles before and after stenting. *Cardiovascular journal of Africa.* 2012;23(9):483-490. doi:10.5830/CVJA-2012-044
22. Cordeiro S, Gomes J, Mendes IC, Martins DS, Sousa A, Anjos R. Doppler Flow Pattern and Arterial Stiffness in Patients with Aortic Coarctation. *Pediatr Cardiol.* 2016;37(8):1465-1468. doi:10.1007/s00246-016-1458-8
23. Wan J, Steele B, Spicer SA, et al. A one-dimensional finite element method for simulation-based medical planning for cardiovascular disease. *Comput Methods Biomech Biomed Engin.* 2002;5(3):195-206. doi:10.1080/10255840290010670

24. Ellwein L, Samyn MM, Danduran M, Schindler-Ivens S, Liebham S, LaDisa JFJ. Toward translating near-infrared spectroscopy oxygen saturation data for the non-invasive prediction of spatial and temporal hemodynamics during exercise. *Biomech Model Mechanobiol.* 2017;16(1):75-96. doi:10.1007/s10237-016-0803-4
25. Westerhof N, Stergiopulos N, Noble MIM. *Snapshots of Hemodynamics*. 1st ed. Springer New York, NY
